# Estimation of COVID-19 spread curves integrating global data and borrowing information

**DOI:** 10.1101/2020.04.23.20077065

**Authors:** Se Yoon Lee, Bowen Lei, Bani K. Mallick

## Abstract

Currently, novel coronavirus disease 2019 (COVID-19) is a big threat to global health. The rapid spread of the virus has created pandemic, and countries all over the world are struggling with a surge in COVID-19 infected cases. There are no drugs or other therapeutics approved by the US Food and Drug Administration to prevent or treat COVID-19: information on the disease is very limited and scattered even if it exists. This motivates the use of data integration, combining data from diverse sources and eliciting useful information with a unified view of them. In this paper, we propose a Bayesian hierarchical model that integrates global data for real-time prediction of infection trajectory for multiple countries. Because the proposed model takes advantage of borrowing information across multiple countries, it outperforms an existing individual country-based model. As fully Bayesian way has been adopted, the model provides a powerful predictive tool endowed with uncertainty quantification. Additionally, the proposed model uses countrywide covariates to adjust infection trajectories in curve fitting. A joint variable selection technique has been integrated into the proposed modeling scheme, which aimed to identify possible country-level risk factors for severe disease due to COVID-19.

## 1 Introduction

Since Thursday, March 26, 2020, the US leads the world in terms of the cumulative number of infected cases for a novel coronavirus, COVID-19. On this day, a dashboard provided by the Center for Systems Science and Engineering (CSSE) at the Johns Hopkins University (https://systems.jhu.edu/-) (Dong et al., 2020) reported that the numbers of the confirmed, death, and recovered from the virus in the US are 83,836, 1,209, and 681, respectively. Figure 1 displays daily infection trajectories describing the cumulative numbers of infected cases for eight countries (US, Russia, UK, Brazil, Germany, China, India, and South Korea), spanning from January 22nd to May 14th, which accounts for 114 days. The dotted vertical lines on the panel mark certain historical dates that will be explained. As seen from the panel, the US has been a late-runner until March 11th in terms of the infected cases, but the growth rate of the cases had suddenly skyrocketed since the day, and eventually excelled the forerunner, China, just in two weeks, on March 26th. Figure 2 shows the cumulative infected cases for 40 countries on May 14th: on the day, the number of cumulative infected cases for the US was 1,417,774 which is more than five times of that of Russia, 252,245.

**Figure 1:**
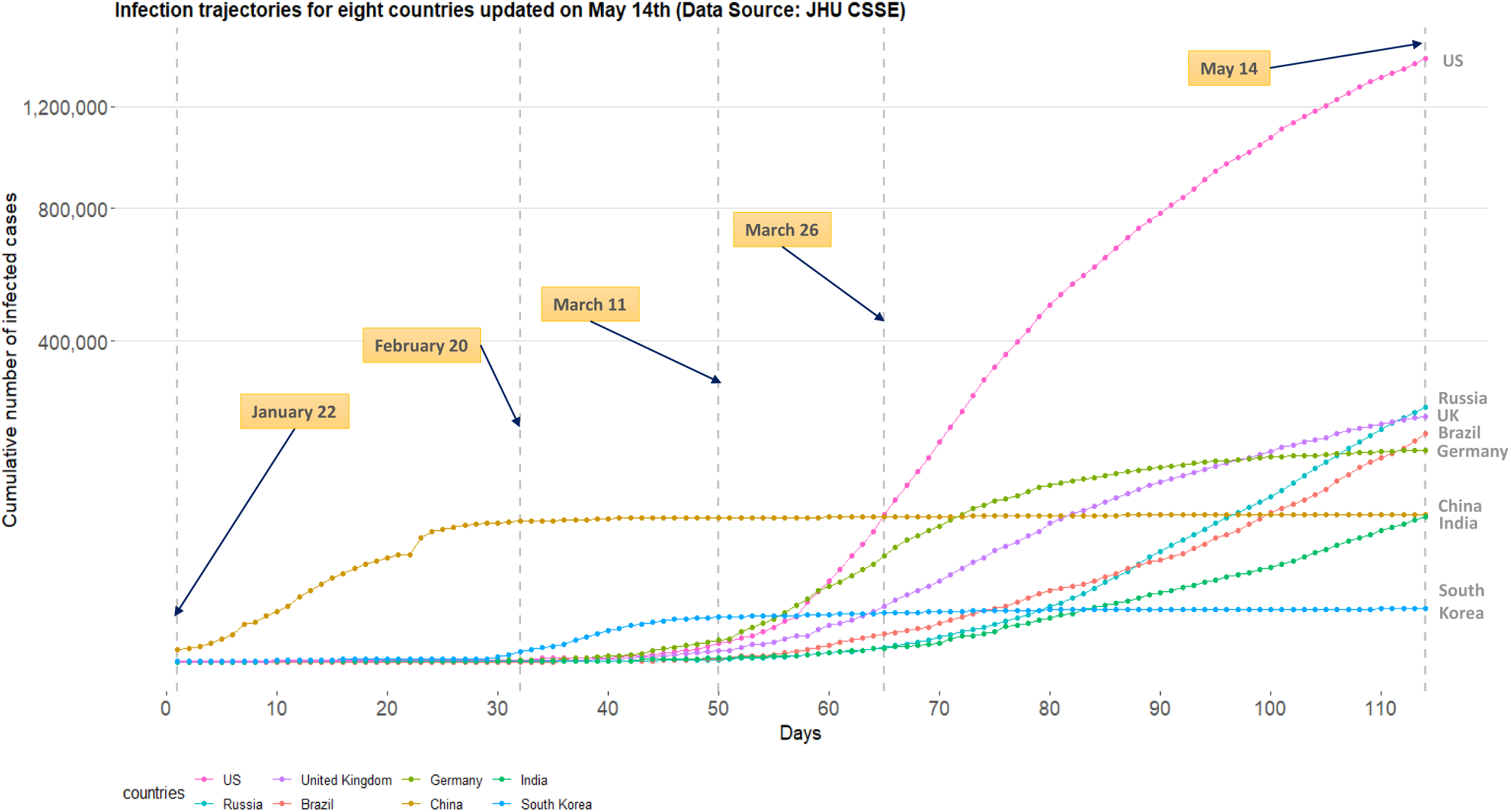
Daily trajectories for cumulative numbers of COVID-19 infections for eight countries (US, Russia, UK, Brazil, Germany, China, India, and South Korea) from January 22nd to May 14th. (Data source: Johns Hopkins University CSSE)

Since the COVID-19 outbreak, there have been numerous research works to better understand the pandemic in different aspects (Gao et al., 2020; Jia et al., 2020; Liu et al., 2020; Peng et al., 2020; Qiang Li, 2020; Remuzzi and Remuzzi, 2020; Sheng Zhang, 2020; Yang et al., 2020). Some of the recent works from statistics community are as follows. Sheng Zhang (2020) focused on a serial interval (the time between successive cases in a chain of transmissions) and used the gamma distribution to study the transmission on Diamond Princess cruise ship. Peng et al. (2020) proposed the generalized susceptible exposed infectious removed model to predict the inflection point for the growth curve, while Yang et al. (2020) modified the proposed model and considered the public health interventions in predicting the trend of COVID-19 in China. Liu et al. (2020) proposed a differential equation prediction model to identify the influence of public policies on the number of patients. Qiang Li (2020) used a symmetrical function and a long tail asymmetric function to analyze the daily infections and deaths in Hubei and other places in China. Remuzzi and Remuzzi (2020) used an exponential model to study the number of infected patients and patients who need intensive care in Italy. One of the major limitations of these works is that the researches are confined by analyzing data from a single country, thereby neglecting the global nature of the pandemic.

One of the major challenges in estimating or predicting an infection trajectory is the heterogeneity of the country populations. It is known that there are four stages of a pandemic: visit economictimes.indiatimes.com/-. The first stage of the pandemic contains data from people with travel history to an already affected country. In stage two, we start to see data from local transmission, people who have brought the virus into the country transmit it to other people. In the third stage, the source of the infection is untraceable. In stage four the spread is practically uncontrollable. In most of the current literature, estimation or prediction of the infection trajectory is based on a single country data where the status of the country falls into one of these four stages. Hence, such estimation or prediction may fail to capture some crucial changes in the shape of the infection trajectory due to a lack of knowledge about the other stages. This motivates the use of data integration (Huttenhower and Troyanskaya, 2006; Lenzerini, 2002) which combines data from different countries and elicits a solution with a unified view of them. This will be particularly useful in the current context of the COVID-19 outbreak.

**Figure 2:**
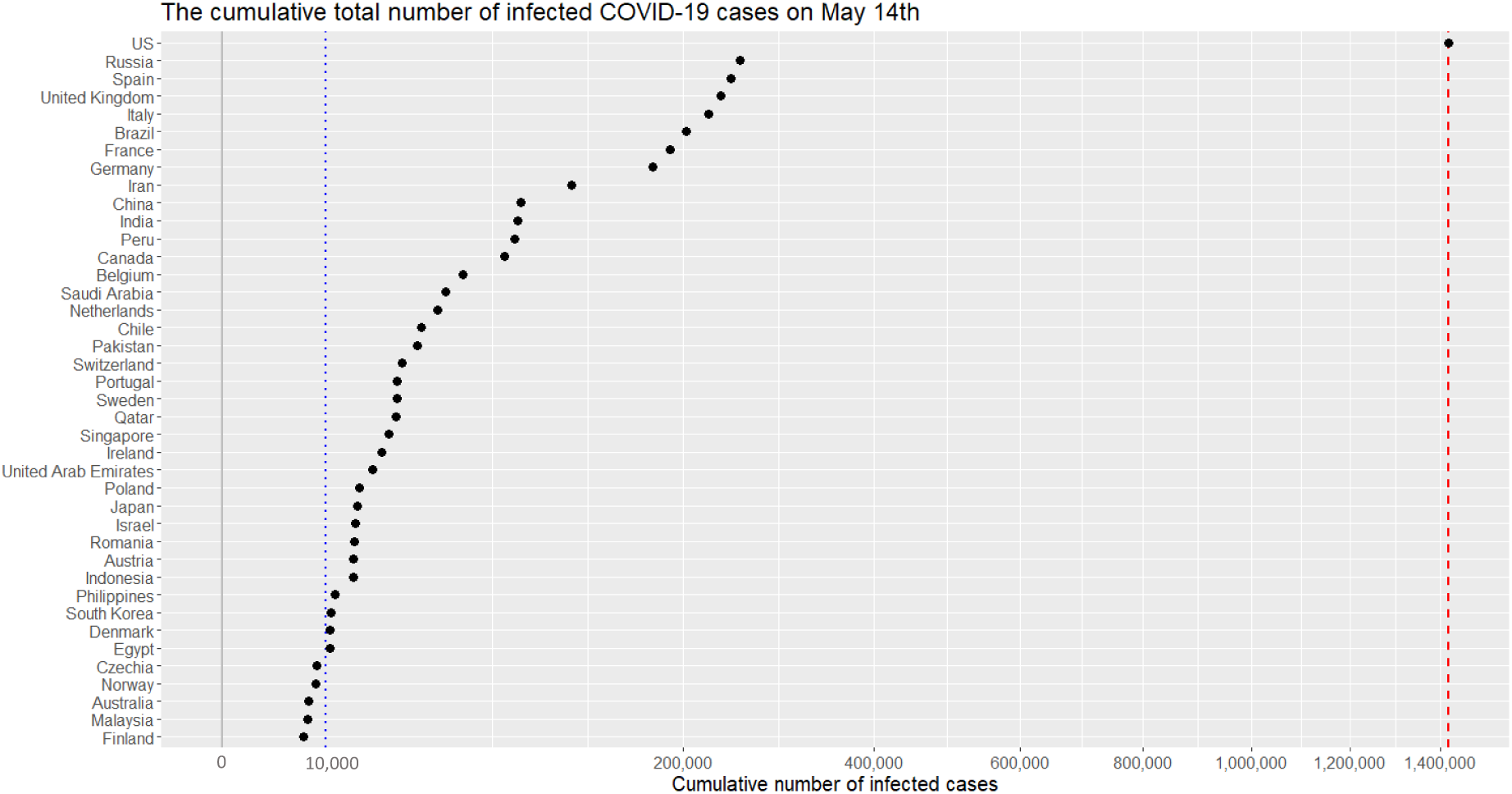
Cumulative numbers of infected cases for 40 countries on May 14th. (*x*-axis are scaled with squared root for visualization purpose.) The red dashed vertical lines represents 1,417,774 cases.

Recently, there are serious discussions all over the world to answer the crucial question: “even though the current pandemic takes place globally due to the same virus, why infection trajectories of different countries are so diverse?” For example, as seen from Figure 1, the US, Italy, and Spain have accumulated infected cases within a short period of time, while China took a much longer time since the onset of the COVID-19 pandemic, leading to different shapes of infection trajectories. It will be interesting to find a common structure in these infection trajectories for multiple countries, and to see how these trajectories are changing around this common structure. Finally, it is significant to identify the major countrywide covariates which make infection trajectories of the countries behave differently in terms of the spread of the disease.

## 2 Significance

The rapid spread of coronavirus has created pandemic, and countries all over the world are struggling with a surge in COVID-19 infected cases. Scientists are working on estimating the infection trajectory for future prediction of cases, which will be useful for future planning and policymaking. We propose a hierarchical model that integrates worldwide data to estimate COVID-19 infection trajectories. Due to information borrowing across multiple countries, the proposed growth curve model will be a powerful predictive tool endowed with uncertainty quantification. Additionally, we use countrywide covariates to adjust curve fitting for the infection trajectory. A joint variable selection technique has been integrated into the modeling scheme, which will identify the possible reasons for diversity among the country-specific infection curves.

## 3 Our Contribution

There are three major classes of infectious disease prediction models: (i) differential equation models, (ii) time series models, and (iii) the statistical models. The differential equation models describe the dynamic behavior of the disease through differential equations allowing the laws of transmission within the population. The popular models include the SI, SIS, SIR, and SEIR models (Hethcote, 2000; Korobeinikov, 2004; Tiberiu Harko, 2014). These models are based on assumptions related to S (susceptible), E (exposed), I (infected), and R (remove) categories of the population. Time series based prediction models such as ARIMA, Grey Model, Markov Chain models have been used to describe dependence structure over of the disease spread over time (Hu et al., 2006; Reza Yaesoubi, 2011; Rushton et al., 2006; Shen X, 2013; Zhirui He, 2018). On the other hand, statistical models, so-called phenomenological models, which follow certain laws of epidemiology (Clayton and Hills, 2013; Thompson et al., 2006) are widely used in real-time forecasting for infection trajectory or size of epidemics in early stages of pandemic (Fineberg and Wilson, 2009; Hsieh, 2009; Pell et al., 2018). Statistical models can be easily extended to the framework of hierarchical models (multilevel models) to analyze data within a nested hierarchy, eventually harnessing the data integration (Browne et al., 2006; Hill, 1965; Stone and Springer, 1965; Tiao and Tan, 1965). In this paper, we use Bayesian hierarchical models so that data integration and uncertainty analysis (Malinverno and Briggs, 2004) are possible in a unified way.

Specifically, we use the Richards growth curve model (Richards, 1959). The novelties of our method are as follows: we (i) use a flexible hierarchical growth curve model to global COVID-19 data, (ii) integrate information from multiple countries for estimation and prediction purposes, (iii) adjust for country-specific covariates, and (iv) perform covariate selection to identify the important reasons to explain the differences among the country-wise infection trajectories. We demonstrate that our proposed models perform better than an individual country-based model.

### 3.1 Richards growth curve models

Richards growth curve model (Richards, 1959), so-called the generalized logistic curve (Nelder, 1962), is a growth curve model for population studies in situations where growth is not symmetrical about the point of inflection (Anton and Herr, 1988; Seber and Wild, 2003). The curve was widely used to describe various biological processes (Werker and Jaggard, 1997), but recently adapted in epidemiology for real-time prediction of outbreak of diseases; examples include SARS (Hsieh, 2009; Hsieh et al., 2004), dengue fever (Hsieh and Chen, 2009; Hsieh and Ma, 2009), pandemic influenza H1N1 (Hsieh, 2010), and COVID-19 outbreak (Wu et al., 2020).

There are variant reparamerized forms of the Richards curve in the literature (Birch, 1999; Cao et al., 2019; Causton, 1969; Kahm et al., 2010), and we shall use the following form in this research

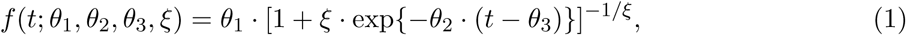

where *θ*_1_, *θ*_2_, and *θ*_3_ are real numbers, and *ξ* is a positive real number. The utility of the Richards curve (1) is its ability to describe a variety of growing processes, endowed with strong flexibility due to the shape parameter *ξ* (Birch, 1999): analytically, the Richards curve (1) (i) becomes the logistic growth curve (Tsoularis and Wallace, 2002) when *ξ* = 1, and (ii) converges to Gompertz growth curve (Gompertz, 1825) as the *ξ* converges to zero from positive side of real numbers. (Gompertz curve is *g*(*t*; *θ*_1_, *θ*_2_, *θ*_3_) = *θ*_1_ · exp [− exp {−*θ*_2_ · (*t* − *θ*_3_)}].) But it is also known that estimation of *ξ* is a complicated problem (Wang et al., 2016), and we resort to a modern sampling scheme, elliptical slice sampler (Murray et al., 2010), to estimate the *ξ*. (See SI Appendix for more detail.)

Figure 3 illustrates roles of the four parameters of the Richards curve (1). The curves on left panel is obtained when (*θ*_1_, *θ*_2_, *θ*_3_) = (10000,0.2,40), while varying the *ξ* to be 1 × 10^−13^(≈ 0), 0.5, and 1, respectively. The right panel pictorially describes the roles of (*θ*_1_, *θ*_2_, *θ*_3_): *θ*_1_ represents the asymptote of the curve; *θ*_2_ is related to a growth rate (analytically, the derivative of logarithm of the curve (1) at *t* = *θ*_3_ is *θ*_2_/2.); and *θ*_3_ sets the displacement along the x-axis. (For more technical detail for the parameters, refer to (Birch, 1999).)

**Figure 3:**
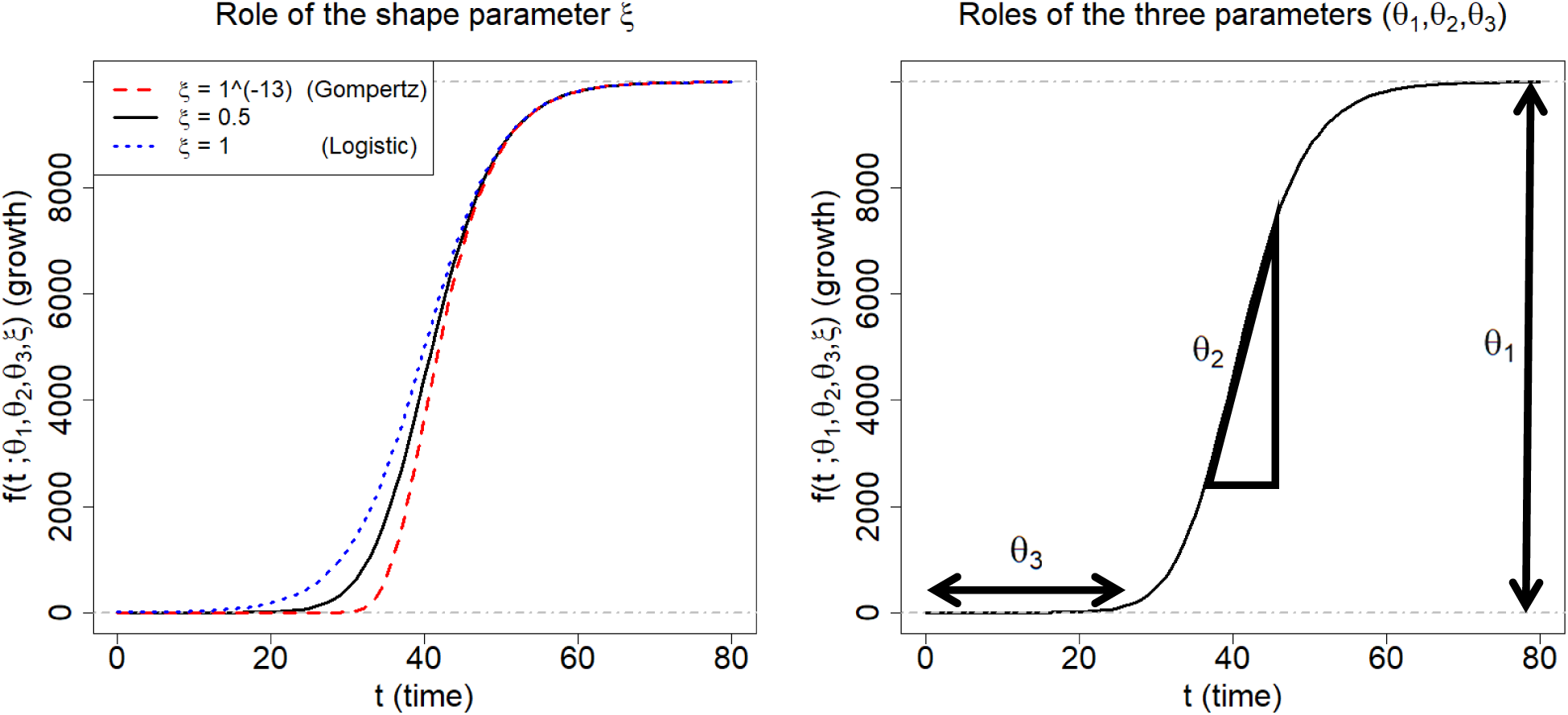
Description of the Richards growth curve model. The curve is obtained when *(θ***_1_***, θ*_2_*, θ*_3_) = (10000, 0.2,40). The left panel is obtained by changing the ξ to be 1 × 10^−13^, 0.5, and 1, respectively. The right panel describes the roles the three parameters in epidemiological modeling: *θ*_1_ represents final epidemic size; *θ*_2_ is an infection rate; and *θ*_3_ is a lag phase.

In epidemiological modeling, the Richards curve (1) can be used as a parametric curve describing infection trajectories shown in the Figure 1. In this context, each of the parameters can be interpreted as follows: *θ***_1_** represents the final epidemic size (that is, the maximum cumulative number of infected cases across the times); *θ*_2_ represents infection rate; and *θ***_3_** represents a lag phase of the trajectory. (The shape parameter *ξ* seems to have no clear epidemiological meaning (Wang et al., 2012).) We shall revisit more detailed interpretations of the parameter in Subsection 4.5.

## 4 Results

### 4.1 Benefits from the information borrowing

We investigate the predictive performance of three Bayesian models based on the Richards growth curve. We start with the individual country-based model (here we use only the single country data) which has been widely used in the literature (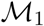). Next, we extend the previous model to a hierarchical model by utilizing the infection trajectories of all the 40 countries (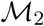). A limitation of 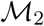 is that it lacks certain countrywide adjustments in estimating the trajectories where the borrowing information takes place uniformly across all the countries although those countries are heterogeneous in terms of aspects like socioeconomic, health environment, etc.. Next, we further upgrade this model by adding country-specific covariates in a hierarchical fashion (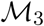). (For technical description for the three models, see the Subsection 6.3.) Eventually, borrowing information across the 40 countries takes place in these two hierarchical models, 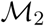 and 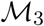, but not in the individual country-based model 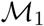.

For evaluation criteria, we calculate the mean squared error (MSE) (Wasserman, 2013) associated with the extrapolated infection trajectory for each of the 40 countries. Training and test data are designated as follows: given that ***y****_k_* = (*y_k_*,_1_,…, *y_k_,_T_*)^┬^ is an infection trajectory of the *k*-th country spanning for *T* days since January 22nd, and *d* is the chosen test-day, then (i) the training data is set by the trajectory spanning for *T* − *d* days since January 22nd (that is, (*y_k_*,_1_,…, *y_k_,_T−d_*)), and (ii) the test data is set by the d recent observations (that is, (*y_k_,_T−d_*_+1_,…,*y_k_,_T_*)).

For the two hierarchical models 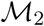 and 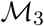, the MSE is averaged over the 40 countries:

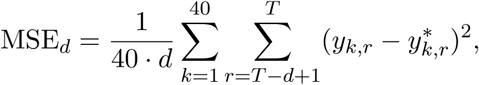

where *y_k_,_r_* is the actual value for the cumulative confirmed cases of the *k*-th country at the *r*-th time point, and 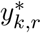 is the forecast value: more concretely, 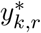 is the posterior predictive mean given the information from 40 countries. For the non-hierarchical model 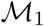, the 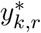 in the MSE*_d_* is acquired by using the data from the *k*-th country.

For each of the short-term test-days (*d* = 5,6,7,8,9,10) and long-term test-days (*d* = 20, 22, 24, 26, 28), we report the median of the MSE*_d_*’s from 20 replicates. The results are shown in Figure 4. From the panel, we see that (1) the predictive performances of two hierarchical models, 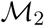 and 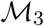, are universally better than that of 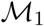 across the number of test-days; and (2) the performance of 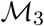 is marginally better than 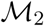. Based on the outcomes, we shall conclude that information borrowing has improved the predictive accuracy in terms of MSE. We present all the results in the consequent subsections based on the model 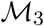. A similar result is found in the *Clemente problem* from (Efron, 2010) where the James-Stein estimator (James and Stein, 1992) better predicts then an individual hitter-based estimator in terms of the total squared prediction error.

**Figure 4:**
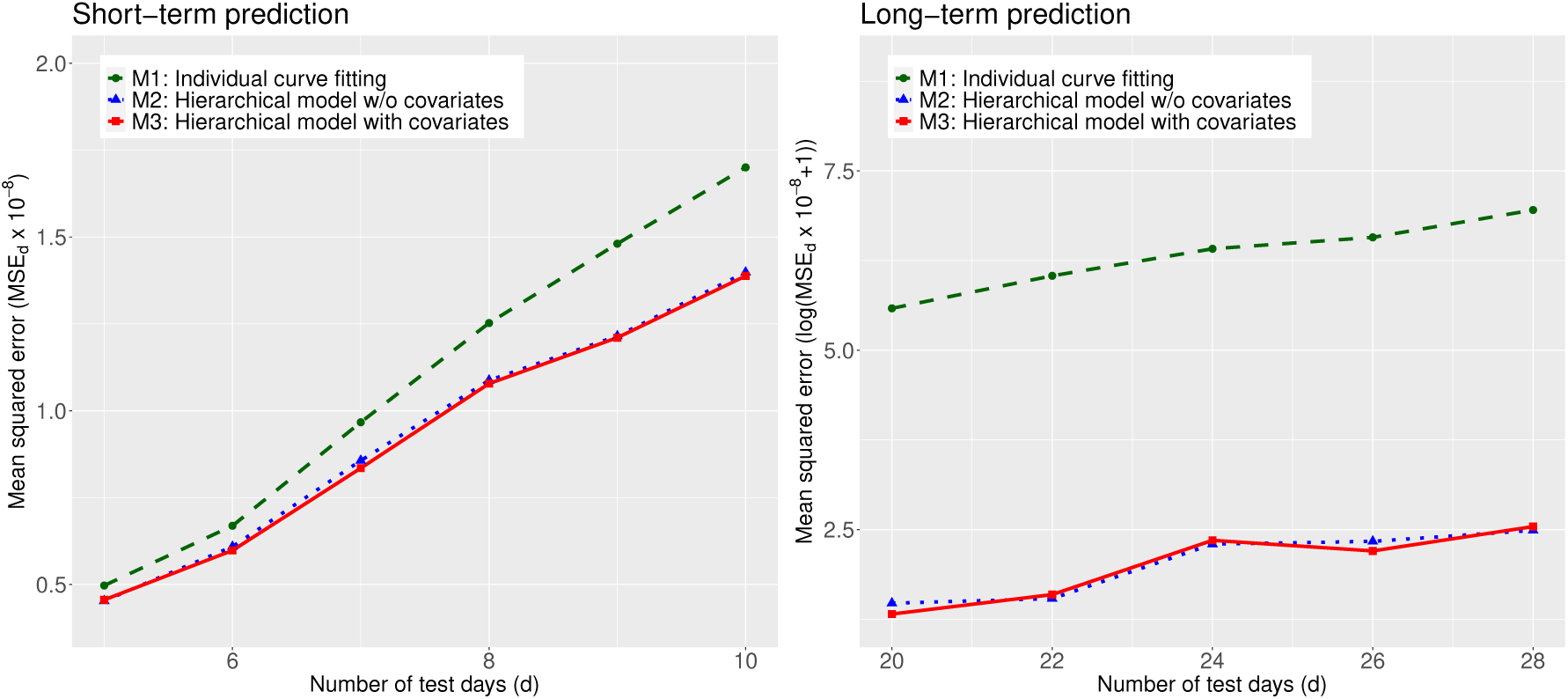
Comparison of the MSE obtained by the three models, 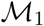, 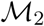, and 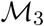, averaged over the 40 countries: short-term (left) and long-term predictions (right). A smaller value for the MSE indicates a better predictive performance.

### 4.2 COVID-19 travel recommendations by country

Centers for Disease Control and Prevention (CDC) categorizes countries into three levels by assessing the risk of COVID-19 transmission, used in travel recommendations by country (Visit www.cdc.gov/-): Level 1, Level 2, and Level 3 indicate the Watch Level (Practice Usual Precautions), Alert Level (Practice Enhanced Precautions), and Warning Level (Avoid Nonessential Travel), respectively.

We categorize the 40 countries into the three levels according to their posterior means for the final epidemic size (that is, *θ*_1_ of the Richards curve (1)). Grouping criteria are as follows: (1) Level 1 (estimated total number is no more than 10,000 cases); (2) Level 2 (estimated total number is between 10,000 and 100,000 cases); and (3) Level 3 (estimated total number is more than 100,000 cases).

Figure 5 displays results of posterior inference for the *θ*_1_ by country, based on the model 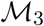. Countries on the *y*-axis are ordered from the severest country (US) to the least severe country (Malaysia) in the magnitude of the posterior means. The red horizontal bars on the panel represent the 95% credible intervals, describing uncertainty for the estimation. Base on the results, there are 14 countries categorized as Level 3 (US, Russia, Brazil, Pakistan, UK, Spain, Italy, India, France, Germany, Peru, Iran, Chile, and Canada). There are 21 countries categorized as Level 2 (from Saudi Arabia to South Korea), and 5 countries categorized as Level 1 (from Czechia to Malaysia).

**Figure 5:**
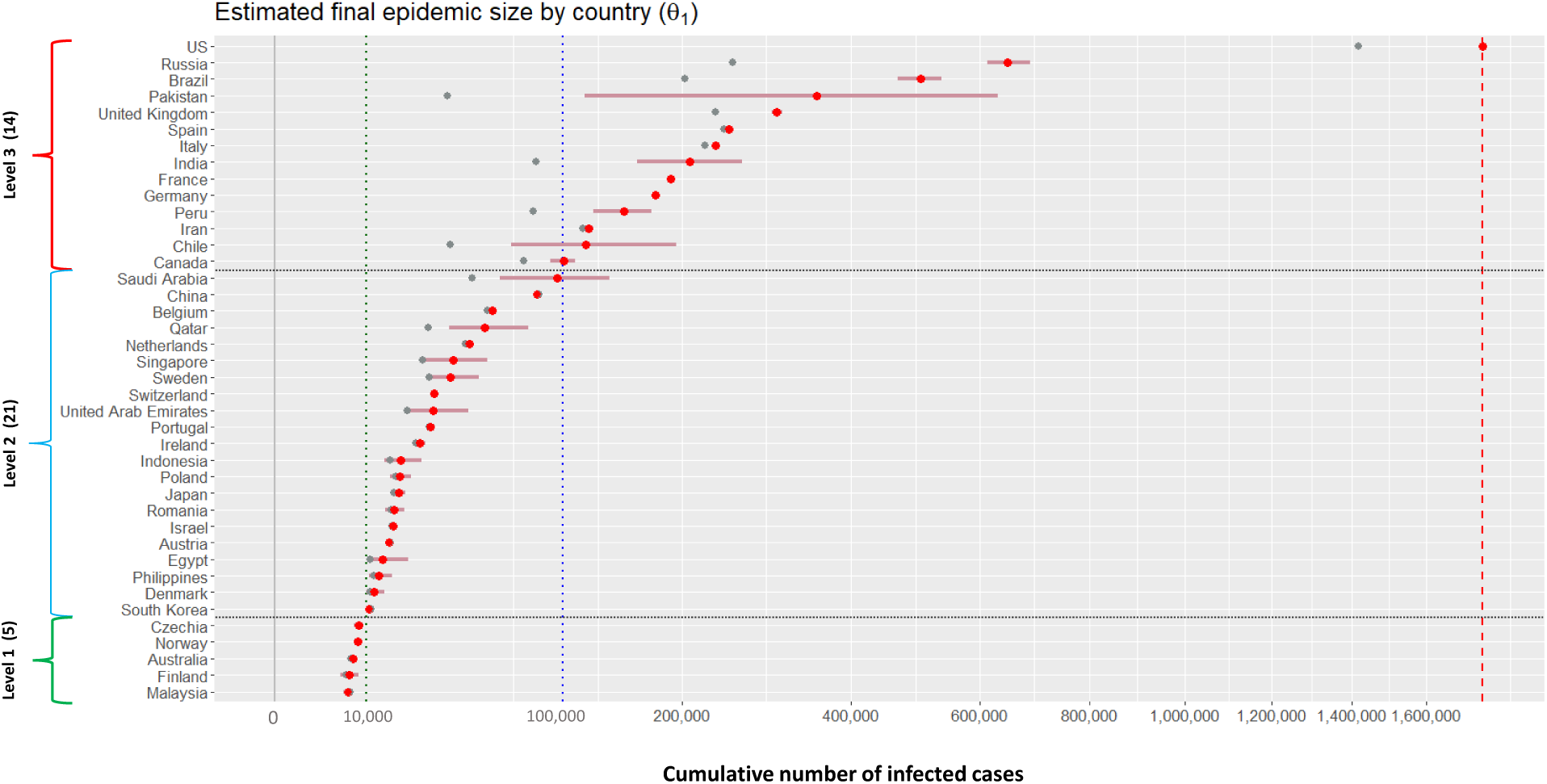
Estimation results for the final epidemic size for 40 countries. Grey dots (•) represent the cumulative numbers of infected cases for 40 countries on May 14th; red dots (•) and horizontal bars (−) represent the posterior means and 95% credible intervals for the *θ*_1_ of the 40 countries. Vertical red dotted line indicates the 1, 760, 569 cases, the posterior mean for the US.

**Figure 6:**
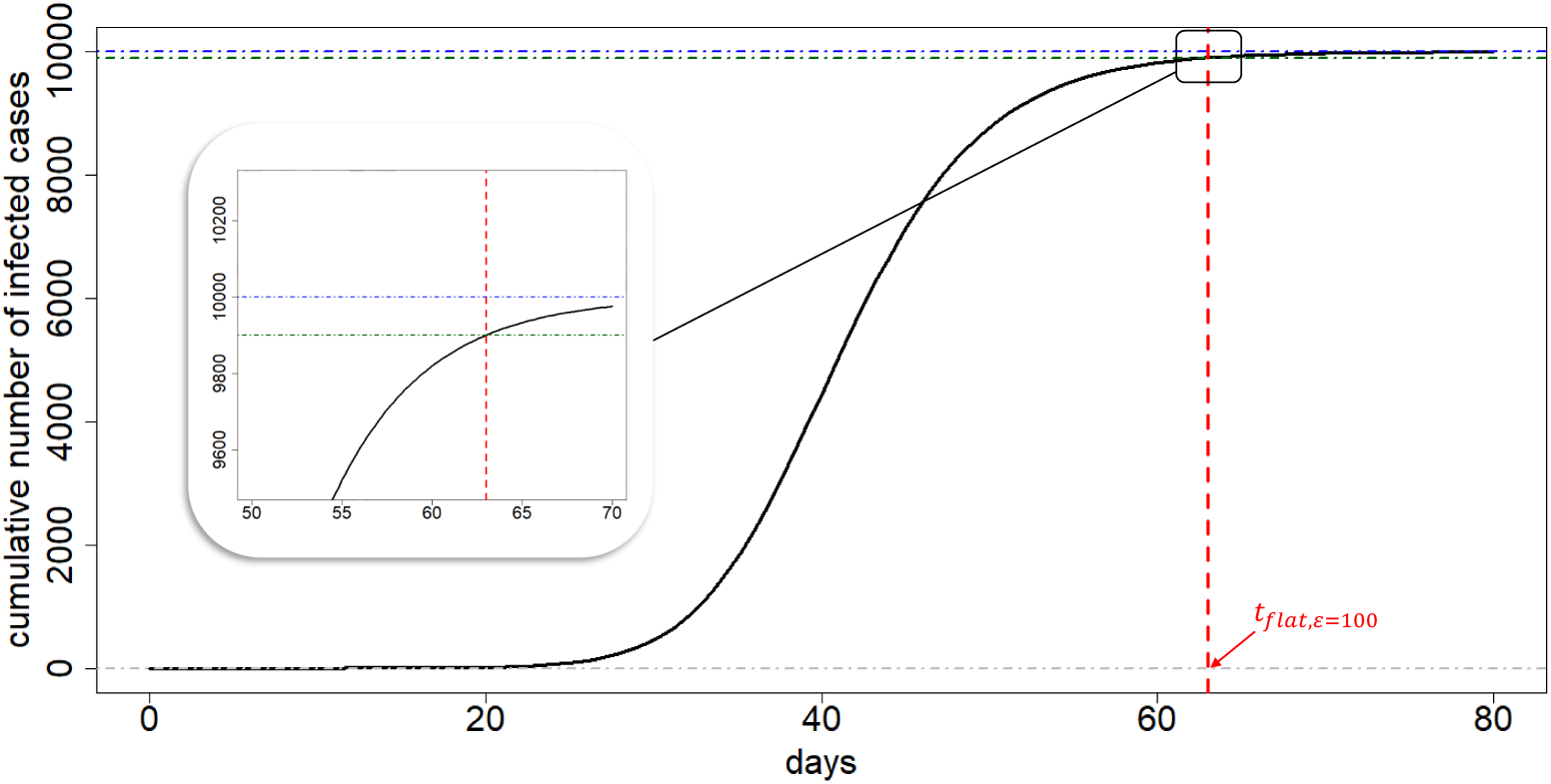
Illustration of flat time point. The exemplary infection trajectory is obtained by the Richards curve when (*θ*_1_, *θ*_2_ *θ*_3_, *ξ*) = (10000, 0.2, 40, 0.5). A flat time point *t*_flat,_*_ϵ_* is approximately 63 (vertical red dashed line). The vertical difference between the *θ*_1_ and the function value of Richards curve evaluated at *t*_flat,ϵ_ is *ϵ* = 100 (cases).

### 4.3 Extrapolated infection trajectories and flat time points

Figure 7 displays the extrapolated infection trajectory (posterior mean for the Richards curve (1)) for the US. The posterior mean of the final epidemic size is 1,760,569 cases. The scenario that ‘millions’ of Americans could be infected was also warned by a leading expert in infectious diseases (Visit a related news article www.bbc.com/-). It is known that prediction of an epidemic trend from limited data during early stages of the epidemic is often futile and misleading (Hsieh et al., 2004). Nevertheless, estimation of a possible severity havocked by the COVID-19 outbreak is an important task when considering the seriousness of the current pandemic situation.

A crucial question is when this trajectory gets flattened. To that end, we approximate a time point where an infection trajectory levels off its value, showing a flattening pattern after the time point. The following is the definition of the *flat time point* which we use in this paper:

#### Definition 4.1

Given the Richards curve *f*(t; *θ*_1_, *θ*_2_, *θ*_3_, *ξ*) (1) and some small *ϵ* > 0, the *flat time point t*_flat,ϵ_ is defined as the solution of the equation *θ*_1_ − *ϵ* = *f*(*t*; *θ*_1_, *θ*_2_, *θ*_3_, *ξ*):

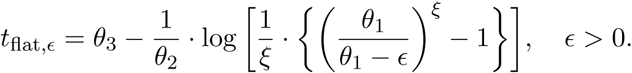

**Figure 7:**
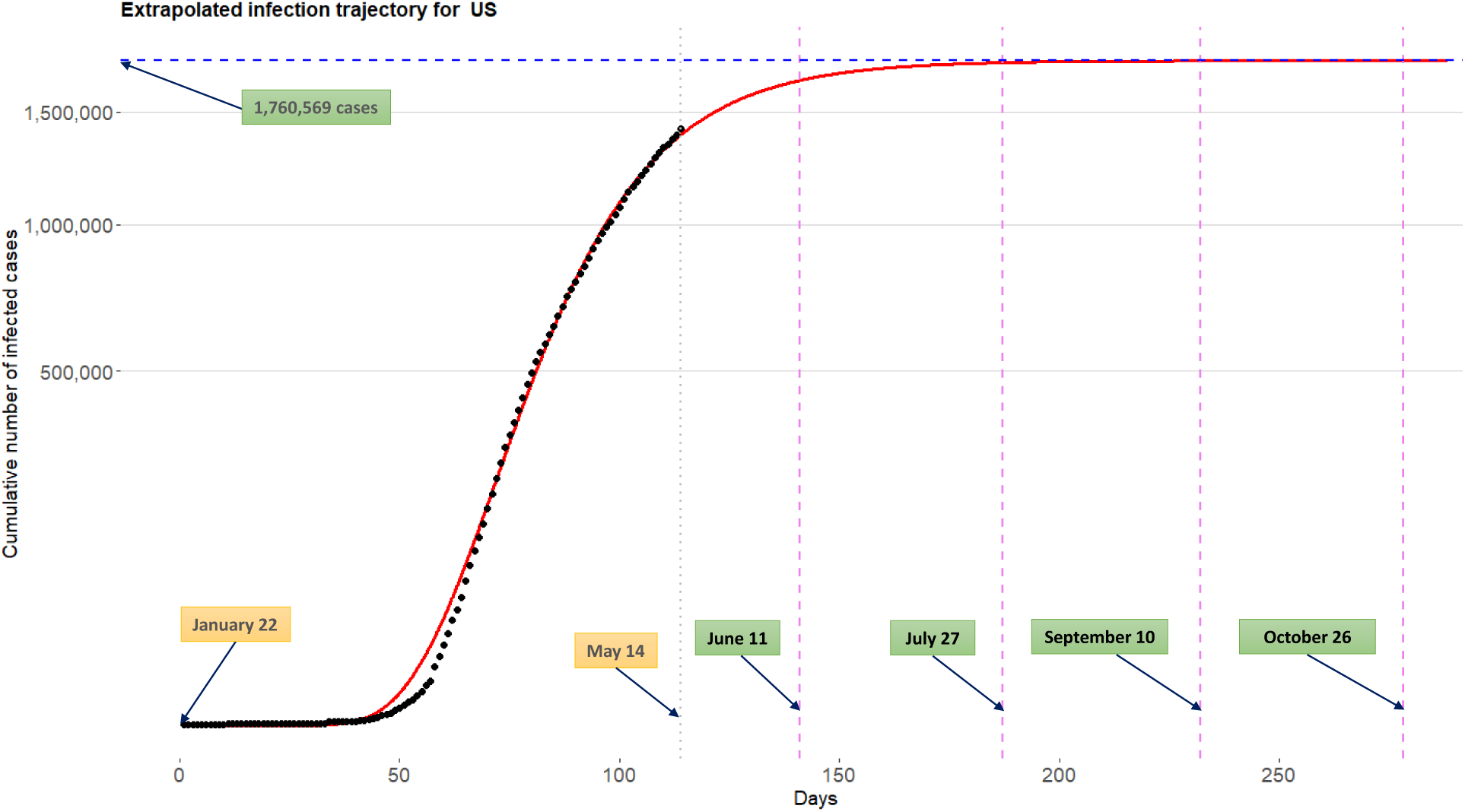
Extrapolated infection trajectory for the US based on the model 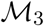. Posterior mean of the maximum number of cumulative infected cases is 1,474,526 cases. Posterior means for the flat time points are *t*_flat,_*_ϵ_*_=100,000_=May 26th, *t*_flat,_*_ϵ_*_=10,000_=July 4th, *t*_flat,_*_ϵ_*_=1,000_=August 11th, and *t*_flat,_*_ϵ_*_=100_=September 18th.

Specifically speaking, the flat time point *t*_flat,_*_ϵ_* is the time point whereat only *ϵ* number of infected cases can maximally take place to reach the final epidemic size *θ*_1_, after the time point *t*_flat,_*_ϵ_*. Figure 6 depicts an exemplary infection trajectory obtained by the Richards curve (1) with (*θ*_1_, *θ*_2_, *θ*_3_, *ξ*) = (10000, 0.2, 40, 0.5). In this case, a flat time point *t*_flat,_*_ϵ_* is approximately 63 when *ϵ* = 100. The choice of *ϵ* > 0 depends on the situation of a country considered: for China which already shows flattening phase (refer to Figure 1) in the infection trajectory, *ϵ* = 1 (case) can be safely used, but for US one may use *ϵ* = 1,000 (cases) or larger numbers.

For the US, the posterior means of the flat time points *t*_flat_,*_ϵ_* are June 11th, July 27th, September 10th, and October 26th when corresponding *ϵ*’s are chosen by 100,000, 10,000, 1,000, and 100, respectively. It is important to emphasize that the extrapolated infection trajectory is *real-time prediction* of COVID-19 outbreaks (Fineberg and Wilson, 2009; Wang et al., 2012) based on observations tracked until May 14th. Certainly, incorporation of new information such as compliance with social distancing or advances in medical and biological sciences for this disease will change the inference outcomes.

Figure 8 show the extrapolated infection trajectories for Russia, UK, and Brazil. Posterior means of the final epidemic size are as follows: (1) for the Russia, 648,190 cases; (2) for the UK, 303,715 cases; and (3) for the Brazil, 503,271 cases. Extrapolated infection trajectory for the Russia (top), UK (middle), and Brazil (bottom). Flat time points are estimated by: (1) for the Russia, *t*_flat,_*_ϵ_*_=1000,000_=June 18th, *t*_flat,_*_ϵ_*_=10,000_=August 2nd, *t*_flat,_*_ϵ_*_=1,000_=September 15th, and *t*_flat,_*_ϵ_*_=100_=October 29th; (2) for the UK, *t*_flat,_*_ϵ_*_=10,000_=June 25th, *t*_flat,_*_ϵ_*_=1,000_=August 10th, and *t*_flat,_*_ϵ_*_=100_=September 25th; and (3) for the Brazil, *t*_flat,_*_ϵ_*_=100,000_=June 6th, *t*_flat,_*_ϵ_*_=10,000_=July 7th, *t*_flat,_*_ϵ_*_=1,000_=August 6th, and *t*_flat,_*_ϵ_*_=100_=September 4th. Results for other countries are included in the SI Appendix.

**Figure 8:**
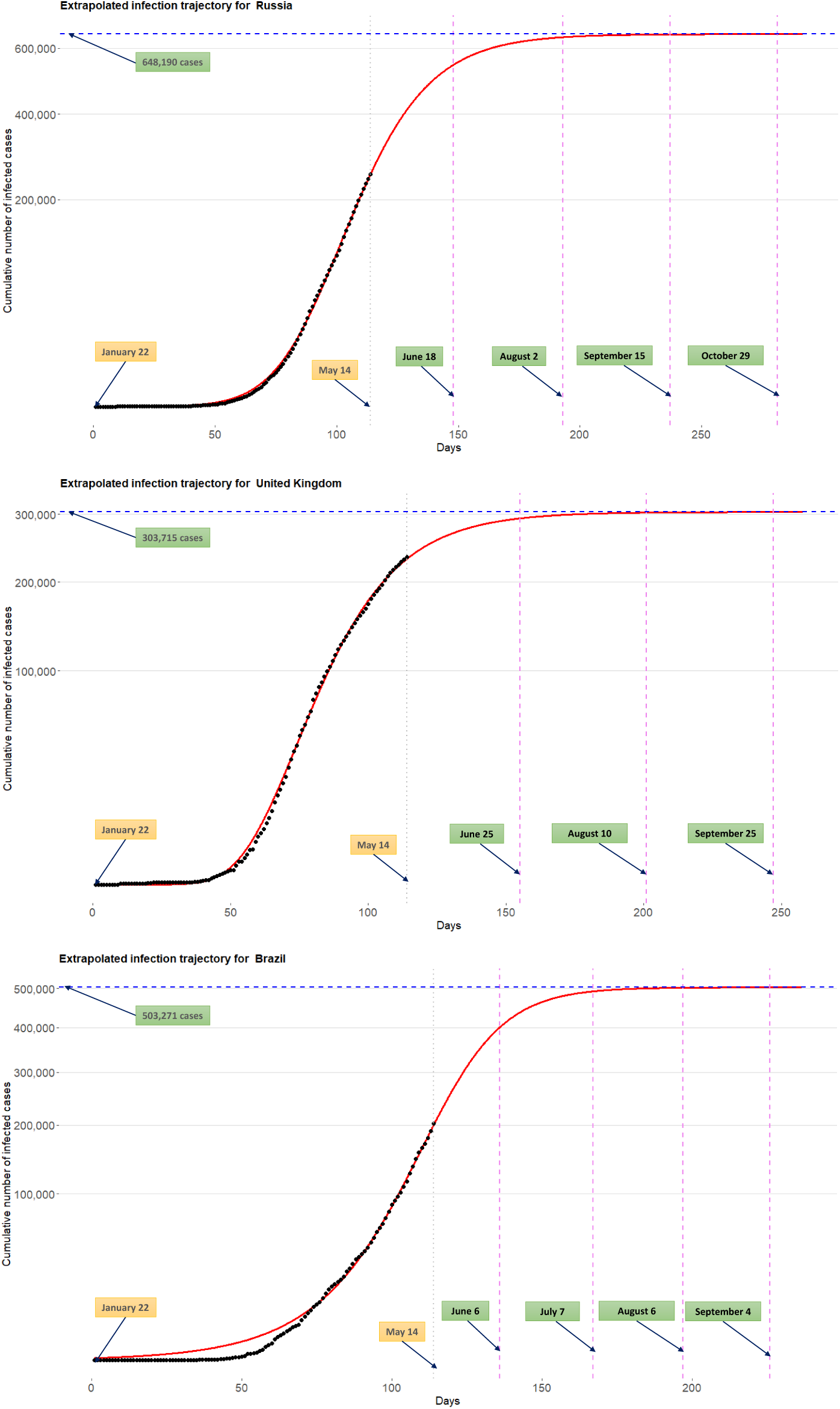
Extrapolated infection trajectory for the Russia (top), UK (middle), and Brazil (bottom). Flat time points are estimated by: (1) for the Russia, *t*_flat,_*_ϵ_*_=100_,_000_=June 18th, *t*_flat,_*_ϵ=_*_10,000_=August 2nd, *t*_flat,_*_ϵ=_*_1,000_=September 15th, and *t*_flat,_*_ϵ=_*_100_=October 29th; (2) for the UK, *t*_flat,_*_ϵ=_*_10,000_=June 25th, *t*_flat,_*_ϵ=_*_1,000_=August 10th, and *t*_flat,_*_ϵ=_*_100_=September 25th; and (3) for the Brazil, *t*_flat,_*_ϵ=_*_100,000_=June 6th, *t*_flat,_*_ϵ=_*_10,000_=July 7th, *t*_flat,_*_ϵ=_*_1,000_=August 6th, and *t*_flat,_*_ϵ=_*_100_=September 4th.

### 4.4 Global trend for the COVID-19 outbreak

Figure 9 displays the extrapolated infection trajectory for grand average over 40 countries obtained from the model 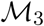. Technically, this curve is acquired by extrapolating the Richards curve by using the intercept terms in linear regressions (3). The grey dots on the panel are historical infection trajectories for 40 countries. Posterior means for the final epidemic size is 145,497 cases. Posterior means for the flat time points are *t*_flat,_*_ϵ_*_=1,000_=June 12th and *t*_flat,_*_ϵ_*_=100_=July 9th.

**Figure 9:**
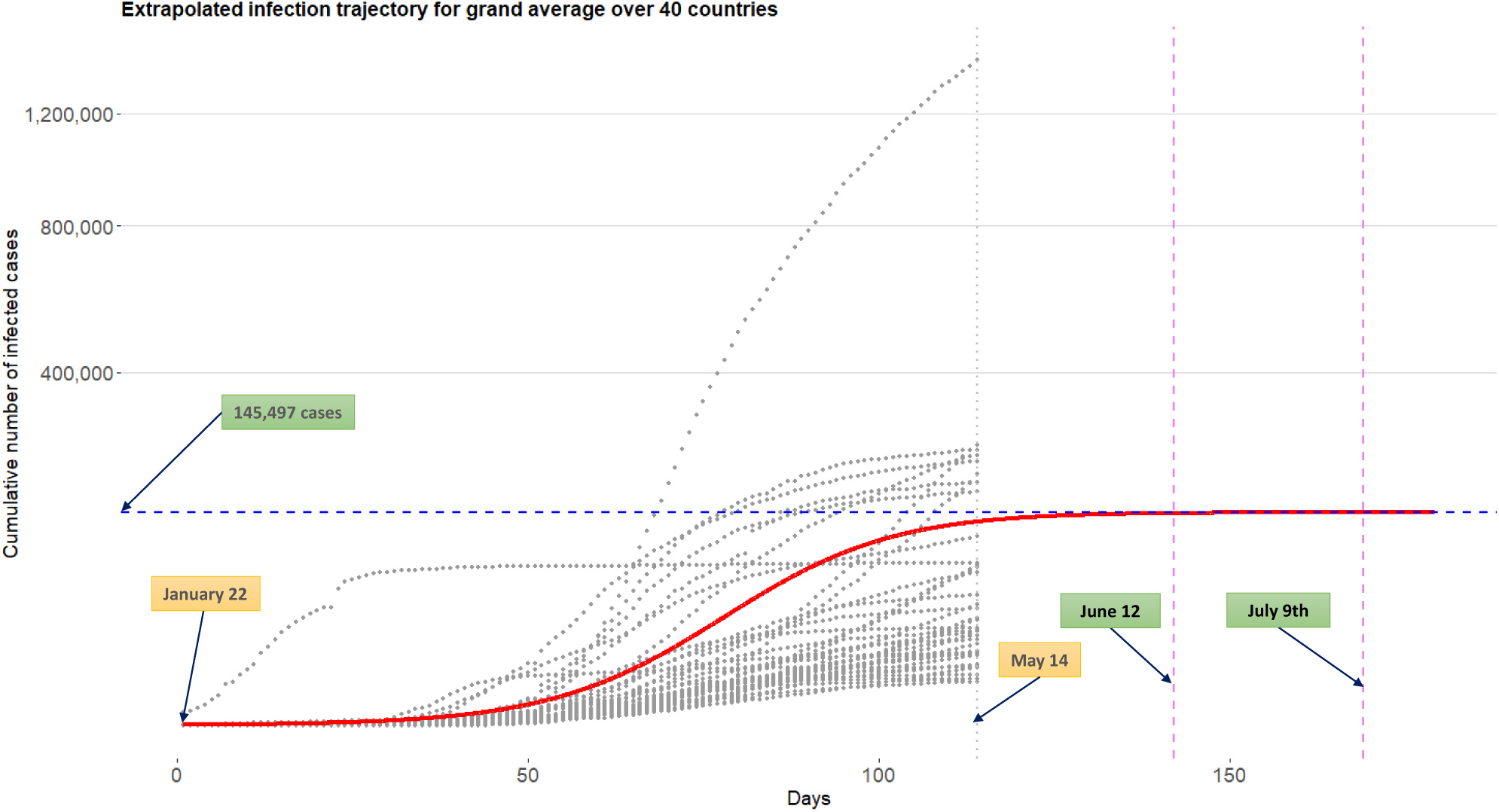
Extrapolated infection trajectory for grand average over 40 countries obtained from the model 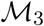. Grey dots are historical infection trajectories for 40 countries spanning from January 22nd to May 14th. Posterior means for the flat time points are *t*_flat,_*_ϵ_*_=1,000_=June 12th and *t*_flat,_*_ϵ_*_=100_=July 9th.

### 4.5 Identifying risk factors for severe disease due to COVID-19

COVID-19 is a new disease and there is very limited information regarding risk factors for this severe disease. There is no vaccine aimed to prevent the transmission of the disease because there is no specific antiviral agent is available. It is very important to find risk factors relevant to the disease. Reliable and early risk assessment of a developing infectious disease outbreak allow for policymakers to make swift and well-informed decisions that would be needed to ensure epidemic control.

CDC described High-Risk Conditions based on currently available information and clinical expertise (For more detail, visit www.cdc.gov/-): those at higher risk for infection, severe illness, and poorer outcomes from COVID-19 include

- People 65 years and older;
- People who live in a nursing home or long-term care facility;
- People with chronic lung disease or moderate to severe asthma;
- People who are immunocompromised, possibly caused by cancer treatment, smoking, bone marrow or organ transplantation, immune deficiencies, poorly controlled HIV or AIDS, and prolonged use of corticosteroids and other immune weakening medications;
- People with severe obesity (body mass index of 40 or higher);
- People with diabetes;
- People with chronic kidney disease undergoing dialysis;
- People with liver disease.

The model 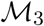 involves three separated linear regressions whose response and coefficient vector are given by *θ_l_* and *β_l_*, respectively (*l* = 1, 2, 3). (See the equation (3)) The sparse horseshoe prior (Carvalho et al., 2009, 2010) is imposed for each of the coefficient vectors which makes the model equipped with covariates analysis. That way, we can identify key predictors explaining the heterogeneity of shapes existing in infection trajectories across 40 countries, and this can be further used in finding risk factors for severe disease due to COVID-19. The results are in table 1 1.

**Table 1:** Important predictors explaining *θ_l_*, *l* = 1, 2, 3

| Final epidemic size ($\theta_1$) | Infection rate ($\theta_2$) | Lag phase ($\theta_3$) |
| --- | --- | --- |
| Insuf_phy_act(+) | Points_of_Entry(-) | Dis_to_China(+) |
| Testing_num_COVID19(+) | Alcohol_consumers_total(+) | Alcohol_cons_rec(-) |
| Testing_popu_COVID19(-) | Alcohol_cons_rec(+) | Median_age(-) |
| Overweight(+) | Air_pollution(+) | Alcohol_consumers_total(-) |
| MCV1_immun(-) | Life_expect_total_60(+) | Testing_num_COVID19(-) |
| Testing_confirm_COVID19(+) | Popu_density(+) | Life_expect_total_60(-) |
| Pol3_immun(-) | Laboratory(-) | Cigarette_smoke(-) |
| Hib3_immun(-) | Heavy_drinking_total(+) | Tuberculosis_case(-) |
| Tempe_avg(-) | Cigarette_smoke(+) | Total_over_65(-) |
| Alcohol_cons_rec(+) | Tobacco_smoke(+) | Tobacco_smoke(-) |

NOTE: the table shows 10 interesting covariates for each parameter. They are listed with “covariate name (sign)” where the sign is taken from the posterior mean for the corresponding coefficient. See SI Appendix for a detailed explanation for the listed covariates.

The followings are general guidelines on how covariates on the Table 1 can be interpreted in the current context of pandemic.

- The parameter *θ*_1_ represents *final epidemic size*. A larger number of *θ*_1_ indicates that a country has (can have) more COVID-19 infected patients in the country. A covariate with plus sign (+) (or minus sign (−)) is a factor associated with an increase (or decrease) of the total infected cases.
- The parameter *θ*_2_ represents *infection rate*. A larger number of *θ*_2_ implies a faster spread of the virus around the country. A covariate with plus sign (+) (or minus sign (−)) is a factor associated with a rapid (or slow) spread of the virus.
- The parameter *θ*_3_ represents *lag phase* of the infection trajectory. The larger the value of *θ*_3_ the later the trajectory begins to accumulate infected cases, leading to a later onset of the accumulation. A covariate with plus sign (+) (or minus sign (−)) is a factor associated with delaying (or bring forward) the onset of the accumulation.

Now, based on the aforementioned guideline, we shall interpret the Table 1 in detail. (The reasoning reflects our subjectivity, and disease expert should decipher precisely.)

As for the parameter *θ*_1_, insufficient physical activity has been selected as one of the important risk factors which may increase the final epidemic size of a country: this implies that certain government policies such as social distancing or remote work system can help decrease the final epidemic size. Additionally, intense immunization coverage on measles, Polio, and Haemophilus Influenzae type B can reduce the final epidemic size. Poor general health status of a population (Jennifer Beam Dowd, 2020) such as overweight and alcohol addiction can increase the epidemic size. (visit related news article www.cidrap.umn.edu/-.) Certain testing information is also associated with the epidemic size, which can be further researched in retrospective studies in swift policymaking for a future pandemic. Finally, the average temperature is negatively related to the epidemic size. (See a WHO report for the relationship between climate change and infectious diseases www.who.int/-.)

Turning to the parameter *θ*_2_, a rigorous fulfillment of general obligations at point of entry is chosen as one of the significant predictors in reducing the infection rate. Additionally, poor smoking and alcoholic behaviors of a country population are risk factors that may increase the infection rate. Demographically, it has been found that densely populated countries or countries where life expectancy is relatively high are more venerable to the rapid disease transmission among people. Among national environmental status, poor air condition which may negatively influence people’s respiratory system is found to be a risk factor increasing the infection rate.

Finally, moving to the parameter *θ*_3_, geological distance from China is an important covariate delaying the onset of the infected cases. The lag of onset is also graphically observed from the Figure 1: time point whereat South Korea begins to accumulate the infected cases is relatively earlier than those of the US, UK, etc. Similar to *θ*_2_, heavier alcohol drinking and tobacco use may result in an earlier onset of the accumulation of the infected patients, thereby bringing forward the infection trajectory. Having larger numbers of median age and elderly people of a population can shorten the lag phase. Finally, conducting frequent testing for the COVID-19 helps detect infected patients, followed by the earlier accumulation for the confirmed cases.

## 5 Discussions

It is important to emphasize that, while medical and biological sciences are on the front lines of beating back COVID-19, the true victory relies on advance and coalition of almost every academic field. However, information about COVID-19 is limited: there are currently no vaccines or other therapeutics approved by the US Food and Drug Administration to prevent or treat COVID-19 (on April 13, 2020). Although numerous research works are progressed by different academic field, the information about COVID-19 is scattered around different disciplines, which truly requires interdisciplinary research to hold off the spread of the disease.

The real-time forecast during early stages of the pandemic may results in premature inference outcomes (Hsieh et al., 2004), but it should not demoralize predictive analysis as the entire human race is currently threatened by unprecedented crisis due to COVID-19 pandemic. To improve the predictive accuracy, data integration from multiple countries is a key notion, which is closely related to borrowing information. The motivation of using the borrowing information is to make use of *indirect evidence* (Efron, 2010) to enhance the predictive performance: for example, to extrapolate the infection trajectory for the US, the information not only from the US *(direct evidence*) but also from other countries *(indirect evidence*) are utilized to better predict the trajectory for the US. Further, to render the information borrowing endowed with uncertainty quantification, Bayesian argument is inevitable, inducing sensible inferences and decisions for users (Lindley, 1972).

The results demonstrated the superiority of our approach compared to an existing individual country-based model. Our research outcomes can be thought even more insightful given that we have not employed information about disease-specific covariates. That being said, using more detailed information such as social mixing data, precise hospital records, or patient-specific information will further improve the performance of our model. Moreover, integration of epidemiological models with these statistical models will be our future topic of research.

## 6 Materials and Methods

### 6.1 Research data

In this research, we analyze global COVID-19 data 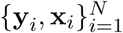, obtained from *N* = 40 countries. (Meanings for the vector notations, **y***_i_* and **x***_i_*, will be explained shortly later.) These countries are most severely affected by the COVID-19 in terms of the confirmed cases on May 14th, and listed on Table 2: each country is contained in the table with format “country name (identifier)”, and this identifier also indicates a severity rank, where a lower value indicates a severer status. The order of the ranks thus coincides with the order of the countries named on the *y*-axis of the Figure 2.

**Table 2:** 40 countries on the research

| Country (index $i$) |
| --- |
| US (1), Russia (2), Spain (3), United Kingdom (4), Italy (5),<br>Brazil (6), France (7), Germany (8), Iran (9), China (10),<br>India (11), Peru (12), Canada (13), Belgium (14), Saudi Arabia (15),<br>Netherlands (16), Chile (17), Pakistan (18), Switzerland (19), Portugal (20),<br>Sweden (21), Qatar (22), Singapore (23), Ireland (24), United Arab Emirates (25),<br>Poland (26), Japan (27), Israel (28), Romania (29), Austria (30),<br>Indonesia (31), Philippines (32), South Korea (33), Denmark (34), Egypt (35),<br>Czechia (36), Norway (37), Australia (38), Malaysia (39), Finland (40) |

NOTE: Countries are listed with the form “country name (identifier)”. This identifier also represents a severity rank. The rank is measured based on the accumulated number of the confirmed cases on May 14th.

For each country *i* (*i* = 1,…, *N*), let *y_it_* denotes the number of accumulated confirmed cases for COVID-19 at the *t*-th time point (*t* = 1,…, *T*). Here, the time indices *t* = 1 and *t* = *T* correspond to the initial and end time points, January 22nd and May 14th, respectively, spanning for *T* = 114 (days). The time series data **y***_i_* = (*y_i_*_1_,…,*y_it_*, …, *y_iT_*)^┬^ is referred to as an *infection trajectory* for the country *i*. Infection trajectories for eight countries (US, Russia, UK, Brazil, Germany, China, India, and South Korea) indexed by *i* = 1, 2, 4, 6, 8, 10, 11, and 33, respectively, are displayed in the Figure 1. We collected the data from the Center for Systems Science and Engineering at the Johns Hopkins University.

For each country *i*, we collected 45 covariates, denoted by x*_i_* = (*x_i_*_1_, …,*x_ij_*,…, *x_ip_*)^┬^ (*p* = 45). The 45 predictors can be further grouped by 6 categories: *the 1st category*: general country and population distribution and statistics; *the 2nd category*: general health care resources; *the 3rd category*: tobacco and alcohol use; *the 4th category*: disease and unhealthy prevalence; *the 5th category*: testing and immunization statistics; and *the 6th category*: international health regulations monitoring. The data sources are the World Bank Data (https://data.worldbank.org/-), World Health Organization Data (https://apps.who.int/-), and National Oceanic and Atmospheric Administration (https://www.noaa.gov/-). Detailed explanations for the covariates are described in SI Appendix.

### 6.2 Bayesian hierarchical Richards model

We propose a Bayesian hierarchical model based on the Richards curve (1), which is referred to as Bayesian hierarchical Richards model (BHRM), to accommodate the COVID-19 data 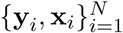. (Although the model is based on the Richards curve, the idea can be generalized to any choice for growth curves.) Ultimately, a principal goal of the BHRM is to establish two functionalities:

a. [Extrapolation] uncover a hidden pattern from the infection trajectory for each country *i*, that is, **y***_i_* = (*y_i_*_1_,…, *y_iT_*) ^┬^, through the Richards growth curve *f*(*t*; *θ*_1_, *θ* _2_, *θ*_3_, *ξ*) (1), and then extrapolate the curve.
b. [Covariates analysis] identify important predictors among the p predictors **x** = (*x*_1_,…, *x_p_*)^┬^ that largely affect on the shape the curve *f*(*t*; *θ*_1_, *θ*_2_, *θ*_3_, *ξ*) in terms of the three curve parameters.

A hierarchical formulation of the BHRM is given as follows. First, we introduce an additive independently identical Gaussian error to each observation 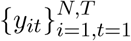, leading to a likelihood part:

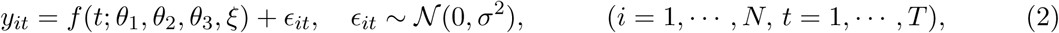

where *f*(*t*; *θ*_1_*_i_*, *θ*_2_*_i_*, *θ*_3_*_i_*, *ξ_i_*) is the Richards growth curve (1) which describes a growth pattern of infection trajectory for the i-th country. Because each of the curve parameters (*θ*_1_, *θ*_2_, *θ*_3_) has its own epidemiological interpretations, we construct three separate linear regressions:

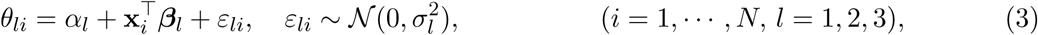

where *β_l_* = (*β_l_*_1_,…,*β_lj_*,…,*β_lp_*)^┬^ is a p-dimensional coefficient vector corresponding to the *l*-th linear regression.

For the shape parameter *ξ*, we assume the standard log-normal prior:

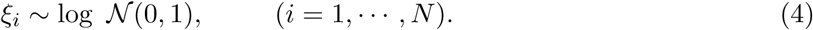

The motivation of choosing the log-normal prior (4) for the *ξ_i_* is that the prior puts effectively enough mass on the region (0, 3) where most of the estimates for the *ξ_i_* (*i* = 1,…, *N*) concentrated on. Additionally, Gaussianity prior assumption makes it possible to employ the elliptical slice sampler (Murray et al., 2010) in sampling from the full conditional posterior distribution of the *ξ_i_*.

To impose a continuous shrinkage effect (Bhadra et al., 2019) on each of the coefficient vectors, we adopt to use the horseshoe prior (Carvalho et al., 2009, 2010):

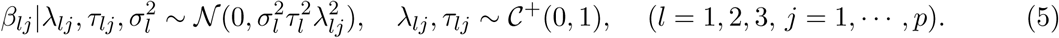

Finally, improper priors (Gelman et al., 2004) are used for the intercept terms and error variances terms in the model:

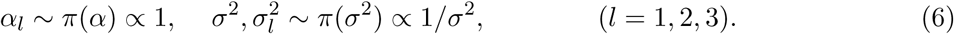

See SI Appendix for a posterior computation for the BHRM (2) – (6).

### 6.3 Technical expressions for three models 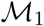, 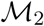, and 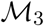

Technical expressions for the three models, 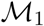, 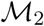, and 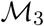, compared in Subsection 4.1 are given as follows:

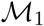 is an individual country-based model (nonhierarchical model) that uses infection trajectory for a single country **y** = (*y*1,…, y_T_)^T^. The model is given by

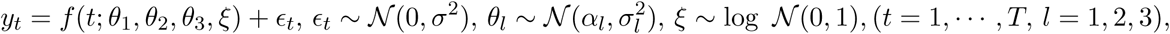

where *f*(*t*; *θ*_1_, *θ*_2_, *θ*_3_) is the Richards growth curve (1), and improper priors (Gelman et al., 2004) are used for error variances and intercept terms as (6).

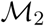 is a Bayesian hierarchical model without using covariates, which uses infection trajectories from *N* countries, 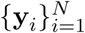. This model is equivalent to BHRM (2) - (6) with removed covariates terms in (3).

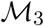 is the BHRM (2) – (6).

## Data Availability

Data used in the research is publicly available.

## Supporting Information Appendix

### Appendix A Tables for covariates

**Table 3:**
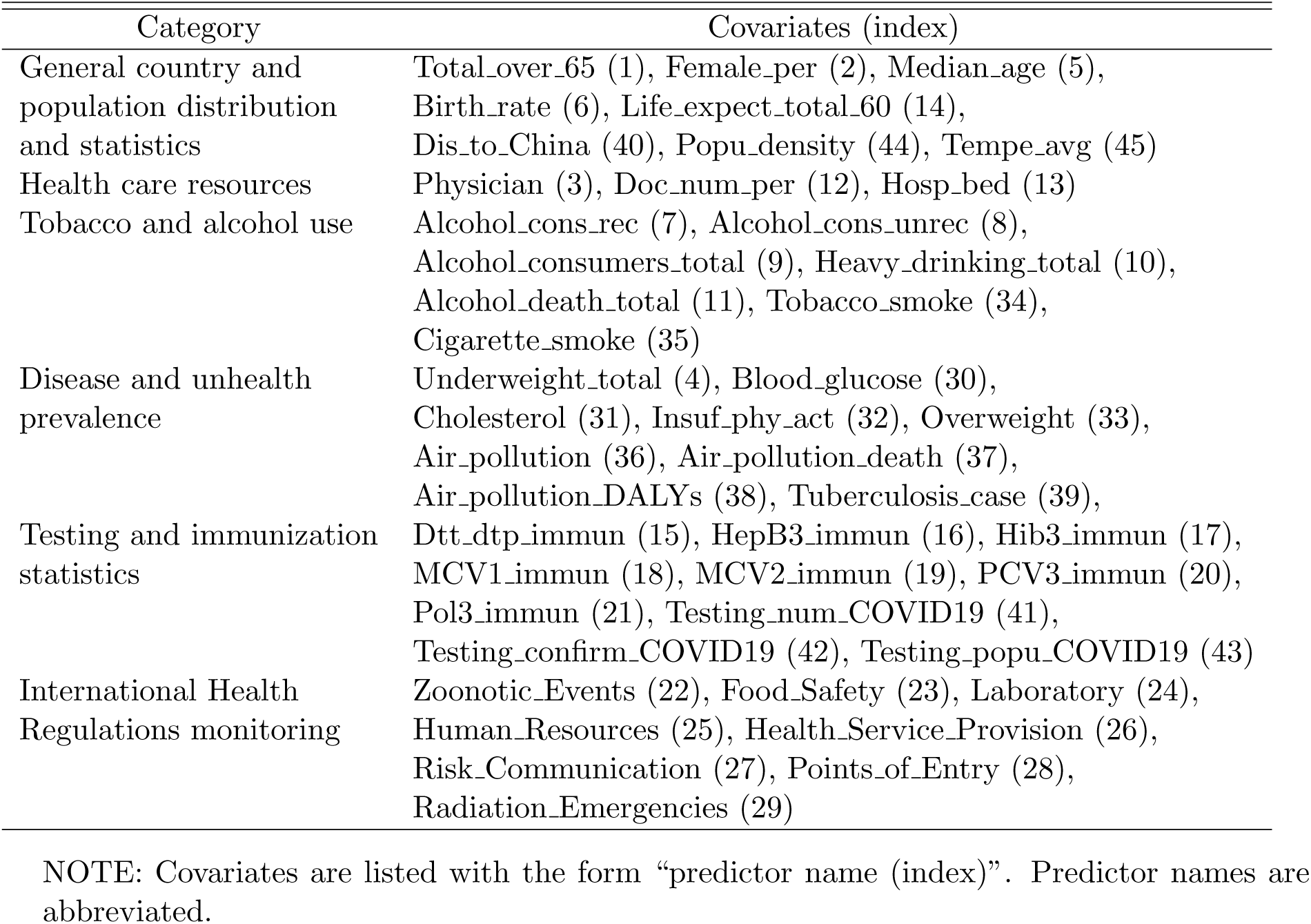
Category of covariates.

**Table 4:** General country and population distribution and statistics.

| Covariates (index $j$) | Explanation |
| --- | --- |
| Total_over_65 (1) | Population ages 65 and above (% of total population) in 2018. |
| Female_per (2) | The percentage of female in the population in 2018. |
| Median_age (5) | Population median age in 2013. |
| Birth_rate (6) | Crude birth rate (per 1000 population) in 2013. |
| Life_expect_total_60 (14) | Life expectancy at age 60 (years) in 2016. |
| Dis_to_China (40) | Calculated by the R function `distm` based on the average longitude and latitude. |
| Popu_density (44) | Population density (people per sq.km of land area) in 2018. |
| Tempe_avg (45) | The average temperature in February and March in the captain of each country (we choose New York for US and Wuhan for China, due to the severe outbreak in the two cities). |

**Table 5:** Health care resources.

| Covariates (index $j$) | Explanation |
| --- | --- |
| Physician (3) | The number of physicians (per 1000 people) between 2015 and 2018. |
| Doc_num_per (12) | The number of medical doctors (per 10000 population) in 2016. |
| Hosp_bed (13) | Average hospital beds (per 10000 population) from 2013 to 2015. |

**Table 6:** Tobacco and alcohol use.

| Covariates (index $j$) | Explanation |
| --- | --- |
| Alcohol_cons_rec (7) | Recorded alcohol consumption per capita (15+) (in litres of pure alcohol), three-year average between 2015 and 2017. |
| Alcohol_cons_unrec (8) | Unrecorded alcohol consumption per capita (15+) (in litres of pure alcohol) in 2016. |
| Alcohol_consumers_total (9) | Alcohol consumers past 12 months (those adults who consumed alcohol in the past 12 months) (% of total) in 2016. |
| Heavy_drinking_total (10) | Age-standardized estimates of the proportion of adults (15+ years) (who have had at least 60 grams or more of pure alcohol on at least one occasion in the past 30 days) in 2016. |
| Alcohol_death_total (11) | Alcohol-attributable death (% of all-cause deaths in total) in 2016. |
| Tobacco_smoke (34) | Age-standardized rates of prevalence estimates for daily smoking of any tobacco in adults (15+ years) in 2013. |
| Cigarette_smoke (35) | Age-standardized rates of prevalence estimates for daily smoking of any cigarette in adults (15+ years) in 2013. |

**Table 7:** Disease and unhealthy prevalence.

| Covariates (index $j$) | Explanation |
| --- | --- |
| Underweight_total (4) | Crude estimate of percent of adults with underweight (BMI < 18.5) in 2016. |
| Blood_glucose (30) | Age-standardized percent of 18+ population with raised fasting blood glucose ($\geq 7.0$ mmol/L or on medication) in 2014. |
| Cholesterol (31) | Percentage of 25+ population with total cholesterol $\geq 240$ mg/dl (6.2 mmol/l) in 2008. |
| Insuf_phy_act (32) | Age-standardized prevalence of insufficient physical activity (% of adults aged 18+) in 2016. |
| Overweight (33) | Age-standardized prevalence of overweight among adults (BMI $\geq 25$) (% of adults aged 18+) in 2016. |
| Air_pollution (36) | Concentrations of fine particulate matter (PM2.5) in 2016. |
| Air_pollution_death (37) | Age-standardized ambient air pollution attributable death rate (per 100000 population) in 2016. |
| Air_pollution_DALYs (38) | Age-standardized ambient air pollution attributable Disability-adjusted life year (DALYs) (per 100000 population) in 2016. |
| Tuberculosis_case (39) | Incidence of tuberculosis (per 100000 population per year) in 2018. |

**Table 8:** Testing and immunization statistics.

| Covariates (index $j$) | Explanation |
| --- | --- |
| Diphtheria tetanus toxoid and pertussis third-dose immunization (15) | Diphtheria tetanus toxoid and pertussis third-dose (DTP3) immunization coverage (% of total 1-year-olds) in 2018. |
| Hepatitis B third-dose immunization (16) | Hepatitis B third-dose (HepB3) immunization coverage (% of total 1-year-olds) in 2018. |
| Haemophilus influenzae type B third-dose immunization (17) | Haemophilus influenzae type B third-dose (Hib3) immunization coverage (% of total 1-year-olds) in 2018. |
| Measles-containing-vaccine first-dose immunization (18) | Measles-containing-vaccine first-dose (MCV1) immunization coverage (% of total 1-year-olds) in 2018. |
| Measles-containing-vaccine second-dose immunization (19) | Measles-containing-vaccine second-dose (MCV2) immunization coverage (% of total nationally recommended age) in 2018. |
| Pneumococcal conjugate vaccines third-dose immunization (20) | Pneumococcal conjugate vaccines third-dose (PCV3) immunization coverage (% of total 1-year-olds) in 2018. |
| Polio third-dose immunization (21) | Polio (Pol3) third-dose immunization coverage (% of total 1-year-olds) in 2018. |
| Testing_num_COVID19 (41) | The number of COVID-19 testing cases (ourworldindata.org/ - collect the data and the data dates are between February and March on several media). |
| Testing_confirm_COVID19 (42) | The total number of confirmed cases on the same day with testing_num divided by the covariate Testing_num_COVID19 (41). |
| Testing_popu_COVID19 (43) | The covariate Testing_num_COVID19 (41) divided by covariate Total_popu (2). |

**Table 9:** International health regulations (IHR) monitoring framework.

| Covariates (index $j$) | Explanation |
| --- | --- |
| Zoonotic_Events (22) | Scores that show whether mechanisms for detecting and responding to zoonoses and potential zoonoses are established and functional in 2018. |
| Food_Safety (23) | Scores that show whether mechanisms are established and functioning for detecting and responding to foodborne disease and food contamination in 2018. |
| Laboratory (24) | Scores that show the availability of laboratory diagnostic and confirmation services to test for priority health threats in 2018. |
| Human_Resources (25) | Scores that show the availability of human resources to implement IHR Core Capacity. |
| Health_Service_Provision (26) | Scores that show an immediate output of the inputs into the health system, such as the health workforce, procurement and supplies, and financing in 2018. |
| Risk_Communication (27) | Scores that show mechanisms for effective risk communication during a public health emergency are established and functioning in 2018. |
| Points_of_Entry (28) | Scores that show whether general obligations at point of entry are fulfilled (including for coordination and communication) to prevent the spread of diseases through international traffic in 2018. |
| Radiation_Emergencies (29) | Scores that show whether mechanisms are established and functioning for detecting and responding to radiological and nuclear emergencies that may constitute a public health event of international concern in 2018. |

NOTE 1: The International health regulations, or IHR (2005), represent an agreement between 196 countries including all WHO Member States to work together for global health security. Through IHR, countries have agreed to build their capacities to detect, assess, and report public health events. WHO plays the coordinating role in IHR and, together with its partners, helps countries to build capacities. (https://www.who.int/ihr/about/-)
NOTE 2: IHR monitoring framework was developed, which represents a consensus among technical experts from WHO Member States, technical institutions, partners and WHO. (https://www.who.int/ihr/procedures/-)

### Appendix B Posterior computation

We illustrate a full description of a posterior computation for the BHRM (2) – (6) by using a Markov chain Monte Carlo (MCMC) simulation (Robert and Casella, 2013). To start with, for illustration purpose, we shall use vectorized notations for the likelihood part (2), regression part (3), and its coe_cients part (5):

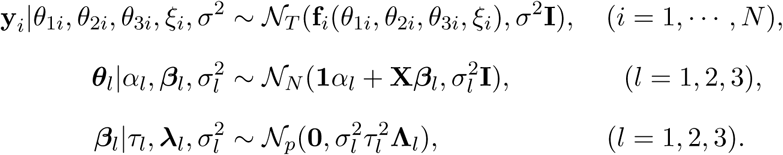

The *T*-dimensional vector *y_i_* = (*y_i_*_1_,…,*y_it_*,…,*yiT*)^┬^ (*i* = 1,…,*N*) is the observed infection trajectory for the country *i* across the times. The notation f(*θ*_1_*_i_*, *θ*_2_*_i_, θ*_3_*_i_, ξ*_i_) (*i* = 1,…,*N*) is *T*-dimensional vector that describes the Richard curves across the times:

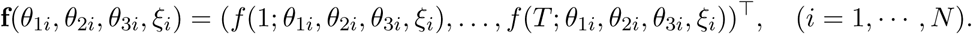

The vectors ***θ****_l_* = (*θ_l_*_1_,…,*θ_lN_*)^┬^ (*l* = 1, 2, 3) and ***ξ*** = (*ξ*_1_,…,*N*)^┬^ are *N*-dimensional vectors for the four parameters of the Richards curve (1) across the *N* countries.

The matrix **X** is *N*-by-*p* design matrix whose *i*-th row vector is given by the *p* predictors x_i_ = (*x_i_*_1_,…,*x_ip_*)^┬^ ∊ *ℝ^p^*, (*i* = 1,…,*N*). The notation **I** stands for an identity matrix. Before implementing, it is recommended that each of column vectors of the design matrix **X** is standardized (Armagan et al., 2013; Tibshirani, 1996): that is, each column vector has been centered, and then columnwisely scaled so that each column vector has mean zero and unit *l*_2_ Euclidean norm.

The *p*-dimensional vector *β_l_* = (*β_l_*_1_,…,*β_lp_*)^┬^ (*l* = 1, 2, 3) denotes *p* coefficients from the *l*-th regression. The vector *λ_l_* = (*λ_l_*_1_,…,*λ_lp_*)^┬^ (*l* = 1, 2, 3) is *p*-dimensional vector for the local-scale parameters, and the matrix **Λ***_l_* is *p*-by-*p* diagonal matrix 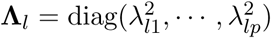 (*l* = 1, 2, 3). The *τ_l_* (*l* = 1; 2; 3) is referred to as the global-scale parameter (Carvalho et al., 2010).

Under the formulation of BHRM (2) – (6), our goal is to sample from the full joint posterior distribution *π*(***θ***_1_, ***θ***_2_, ***θ***_3_, ***ξ***,σ^2^,Ω_1_, Ω_2_, Ω_3_|y_1:_*_N_*) where 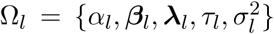 (*l* = 1, 2, 3), whose proportional part is given by

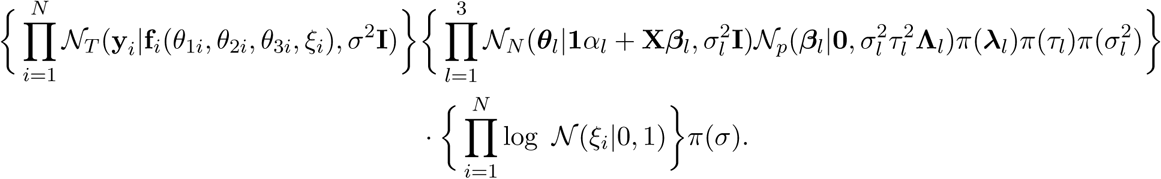

To sample from the full joint density, we use a Gibbs sampler (Casella and George, 1992) to exploit conditional independences among the latent variables induced by the hierarchy. The following algorithm describes a straightforward Gibbs sampler

***Step 1***. Sample ***θ***_1_ from its full conditional distribution

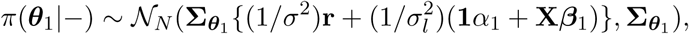

where 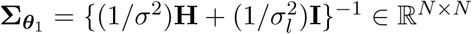. Here, the vector **r** is a *N*-dimensional vector which is given by 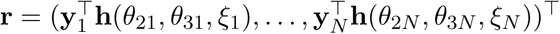 such that the *T*-dimensional vector **h**(*θ*_2i_, *θ*_3_*_i_*, *ξ_i_*) (*i* = 1,…,*N*) is obtained by

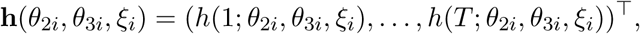

where *h*(*t*; *θ*_2_, *θ*_3_, *ξ*) = [1 + *ξ·*exp{−*θ*_2_·(*t* − *θ*_3_)}]^−1/^*^ξ^*

***Step 2***. Sample *θ*_2_*_i_* and *θ*_3_*_i_*, *i* = 1,⋯,*N*, independently from their full conditional distributions. Proportional parts of the distributions are given by

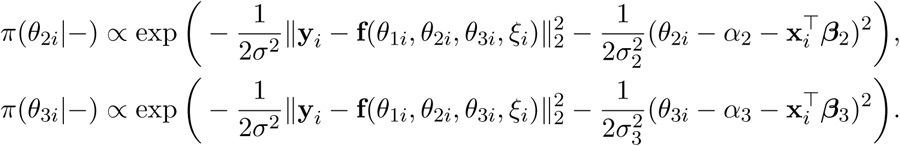

Here, ||·||_2_ indicates the *l*2-norm. Note that the two conditional densities are not known in closed forms because two parameters, *θ*_2_*_i_* and *θ_3i_*, participate to the function *f*(*θ*_1_*_i_*, *θ*_2_*_i_*, *θ*_3_*_i_*, *ξ_i_*) in a nonlinear way. We use the Metropolis algorithm (Andrieu et al., 2003) with Gaussian proposal densities within this Gibbs sampler algorithm.

***Step 3***. Sample *ξ_i_*, *i* = 1,⋯,*N*, independently from its full conditional distribution. Proportional parts of the distributions are given by

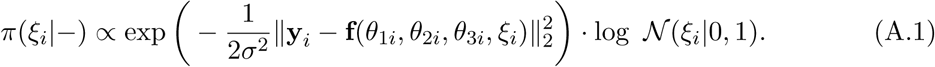

Note that the density (A.1) is not expressed in a closed form distribution. Because the shape parameter *ξ_i_* is supported on (0, ∞) and participates in the Richards curve (1) as an exponent, sampling from the density needs a delicate care, where by we employed the elliptical slice sampler (Murray et al., 2010).

***Step 4***. Sample *σ*_2_ from its full conditional distribution

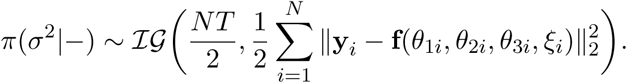

***Step 5***. Sample *α_l_*, *l* = 1, 2, 3, independently from their full conditional distributions

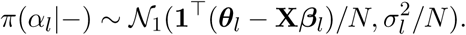

***Step 6***. Sample *β_l_*, *l* = 1, 2, 3, independently from conditionally independent posteriors

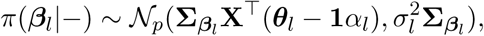

where 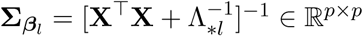, 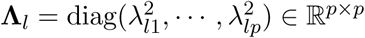, and **Λ**_*_*_l_* = *τ*^2^ ***Λ****_l_*.

***Step 7***. Sample *λ_lj_*, *l* = 1, 2, 3, *j* = 1,⋯,*p*, independently from conditionally independent posteriors

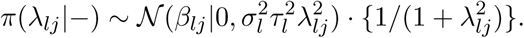

Note that the densities *π*(*λ_lj_* |−) (*l* = 1, 2, 3, *j* = 1,⋯, *p*) are not expressed in closed forms: we use the slice sampler (Neal, 2003).

***Step 8***. Sample *τ_l_*, *l* = 1, 2, 3, independently from conditionally independent posteriors

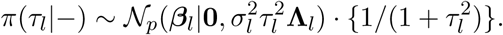

Note that the densities *π*(*τ_l_|−*) (*l* = 1, 2, 3) are not expressed in closed forms: we use the slice sampler (Neal, 2003).

***Step 9***. Sample 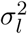, *l* = 1; 2; 3, independently from their full conditionally distributions

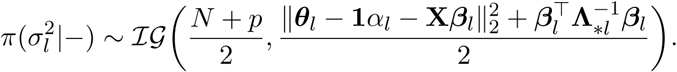

#### B.1 Elliptical slice sampler for Step 3

To start with we shall use the variable change (*η* = log *ξ*) to the right hand side of (A.1):

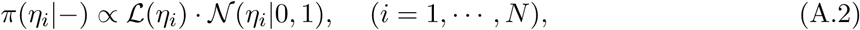

such that 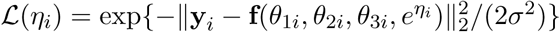 corresponds to a likelihood part.

Now, we use the elliptical slice sampler (ESS) (Murray et al., 2010; Nishihara et al., 2014) to sample from 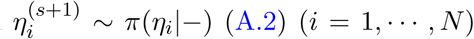 that exploits the Gaussian prior measure. Conceptually, ESS and the Metropolis-Hastings (MH) algorithm (Chib and Greenberg, 1995) are similar: both methods are comprised of two steps: *proposal step and criterion step*. A difference between the two algorithms arises in the *criterion step*. If the new candidate does not pass the criterion, then MH takes the current state as the next state: whereas, ESS re-proposes a new candidate until rejection does not take place, rendering the algorithm rejection-free. Further information for ESS is referred to the original paper (Murray et al., 2010). After ESS has been employed, the realized *η_i_* needs to be transformed back through *ξ* = *e^η^*. Algorithm 1 illustrates the full description of algorithms: to avoid notation clutter, the index *i* is omitted.

##### Algorithm 1

Elliptical slice sampler to sample from *π*(*ξ|−*)

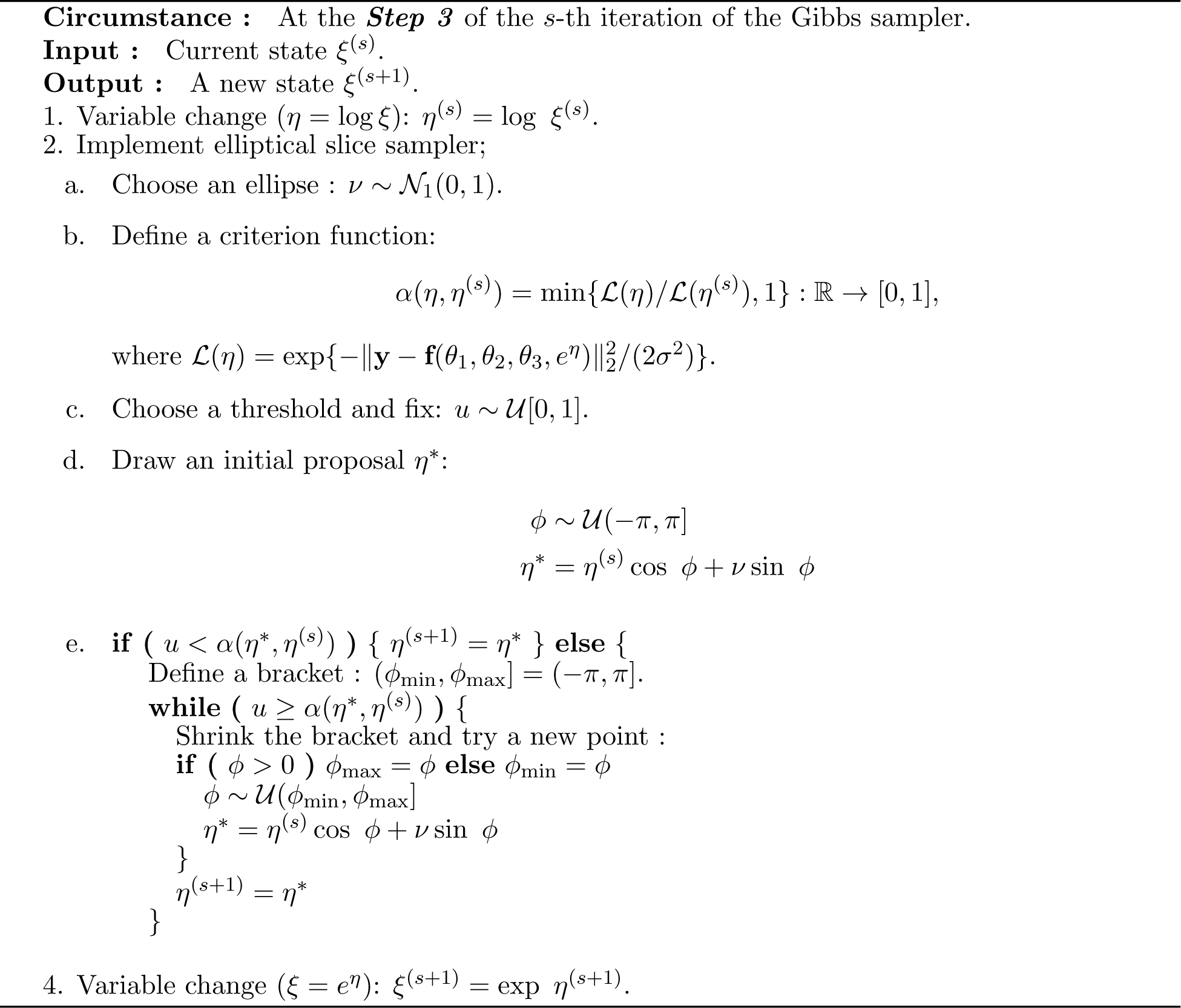

### Appendix C Infection trajectories for the top 20 countries

The file includes extrapolated infection trajectories for the top 20 countries that are most severely affected by the COVID-19.

